# The Clinician Model Card: development and evaluation of clinician-centered documentation for AI-based clinical decision support

**DOI:** 10.64898/2026.04.15.26350930

**Authors:** Louis Agha-Mir-Salim, Nicolas Frey, Zoe Kaiser, Lina Mosch, Eva Weicken, Oscar Freyer, Jackie Ma, Mirja Mittermaier, Alexander Meyer, Stephen Gilbert, Claudia Müller-Birn, Felix Balzer

## Abstract

AI documentation frameworks remain poorly designed for point-of-care use, leaving clinicians without actionable information on how to use clinical AI models when they need it most. We developed the Clinician Model Card, an interactive, clinician-centered documentation tool, and evaluated it in a sequential exploratory mixed-methods study: interviews with 12 physicians informed iterative co-design, evaluated in a national survey of 129 physicians across Germany. The tool was well-received: 84% agreed it should be routinely available, and 66% considered its content relevant to clinical decision-making. Yet comprehensibility of statistical performance metrics remained poor despite targeted interventions: only 32% understood the *Validation & Performance* section well, and fewer than 54% correctly interpreted AUROC or PPV, with AI literacy as strong predictor of comprehension (ρ = 0.59). We propose empirically derived design principles for clinician-centered AI documentation. Effective AI transparency requires not only clinician-friendly design and workflow integration, but sustained investment in AI literacy.

## Introduction

A sepsis prediction model flags a patient in the emergency department as having a 78% probability of developing sepsis within the next four hours. The physician initiates treatment. The system’s intended workflow, however, specifies that a flagged patient should first prompt a team discussion, and the model’s low positive predictive value means most alerts do not indicate true sepsis. The AI output in form of a single number was visible; the context needed to use it appropriately was not.

This scenario highlights a fundamental tension in clinical AI deployment. As AI-based clinical decision support systems (CDSS) enter healthcare^1^, their value depends not only on technical performance but on whether clinicians can interpret and appropriately act on their outputs^2^. Yet clinicians, the primary users, often lack critical information: how a model was developed, for which patients it was validated, how outputs should influence workflow, and how performance metrics apply in a specific clinical encounter^3–6^. This is not merely an educational problem; it reflects a structural gap between AI development outputs and the information clinicians actually receive at the point of care^7^.

Regulatory frameworks and scientific initiatives have begun to address this gap. AI-based CDSS must comply with regulations mandating clear documentation and human oversight, for example, the EU AI Act and comparable requirements enforced by the US FDA and Singapore Health Sciences Authority^8–10^. Complementary scientific initiatives include DECIDE-AI^11^, STARD-AI^12^, and CONSORT-AI^13^ for study-level reporting, and Model Cards^14^ for standardized documentation of AI models. Sendak et al.’s Model Facts Label^15^ collates AI model information for clinicians, and recent contributions, including the CHAI Model Card^16^ and layered documentation proposals^17^, further emphasize timely relevance.

Despite these advances, a critical gap persists between documentation as it exists and documentation as it functions at the point of care. Existing formats were not designed for real-time clinical use. They present technical metrics and statistical terminology in a raw, unfiltered form that is cognitively demanding to parse under time pressure^18,19^. They remain single-layered and non-interactive^20^ and were developed without systematic elicitation of clinician information needs and AI literacy. As a result, the clinician at the bedside confronts either a dense technical document or nothing at all. Despite proposals for user-centered documentation^17^, there is no empirical evidence on what clinicians actually need, how they engage with structured AI documentation in practice, and which design features support or impede comprehension^18^.

To address this gap, we developed and evaluated the Clinician Model Card, an interactive, clinician-centered documentation tool designed for point-of-care use. Using a sequential exploratory mixed-methods design^21^, we first identified clinician information needs and comprehension barriers through semi-structured interviews, translated these into an interactive prototype, and evaluated it in a national survey among physicians. Our aim was not only to produce a tool but to generate empirically derived design principles for clinician-centered AI documentation grounded in what clinicians actually need, struggle with, and prioritize at the moment of clinical decision-making in conjunction with AI.

## Results

The study proceeded in three sequential phases. Qualitative interviews with 12 physicians (Phase 1) identified clinician information needs, comprehension barriers, and accessibility requirements, which informed an iterative co-design process resulting in the Clinician Model Card prototype (Phase 2). The prototype was then evaluated in a national survey of 129 physicians across Germany (Phase 3). Across both phases, a consistent picture emerged: clinicians found the card readable and clinically relevant, and strongly preferred active workflow integration over passive availability. Yet a persistent and striking gap remained: comprehensibility of statistical performance metrics was low despite targeted design interventions, with fewer than half of respondents correctly interpreting key metrics and only 32% reporting they understood the *Validation & Performance* section well. This tension between perceived utility and actual comprehension is a central finding of the study.

### Phase 1: Qualitative interviews

#### Participants

We interviewed 12 physicians (9 male, 3 female) from internal medicine (n = 7) and anesthesiology (n = 5). Ten were residents and two were clinician-scientists. Only one had prior familiarity with the concept of a Model Card. Participant characteristics are summarized in Supplementary Table 1.

#### Relevance and content prioritization

Card sorting of the Model Facts Label^15^ revealed a consistent hierarchy of perceived clinical relevance across participants. Identifying information, the *Summary*, and *Uses and Directions* were ranked highest; *Warnings* were considered more important than their position in the existing Model Facts Label implied; *Validation and Performance* was ranked second last and viewed as largely peripheral to clinical decision-making. This ordering reflected a clear preference for actionable, decision-relevant content over technical documentation. A recurrent and strongly expressed information need was instance-level explainability: participants wanted to understand the rationale behind individual predictions, not only aggregate model statistics. As one participant put it: *“Yes, and then of course there’s the whole topic of explainability. I would definitely question it — or very much want to know how this prognosis, a 90 percent probability, came about, and whether there were any specific factors that were particularly decisive for it“* (P6). The full ranking and code system are provided in Supplementary Tables 2 and 3.

#### Comprehensibility

Sections covering model identity, approval status, summary, mechanism, and use directives were generally considered clear. Statistical terminology, including sensitivity, positive predictive value (PPV), and validation metrics (e.g., AUROC, area under the receiver operating characteristic curve), was consistently described as difficult to interpret. One participant captured a widely shared concern: *“I imagine that for many clinicians who may not be so well versed in statistics, this classification of positive predictive value and sensitivity is probably quite complicated and overwhelming“* (P1). Participants recommended visual aids to represent performance metrics, plain-language explanations of technical terms, and a layered approach providing brief summaries for routine use with more detailed content available on demand.

#### Accessibility

Two distinct contexts of use shaped participants’ access preferences: during an onboarding phase when a new system is introduced, requiring comprehensive engagement, and an on-demand reference function during routine care, triggered when a result appears unexpected or contradictory and requiring rapid, targeted access. Participants consistently emphasized that information must surface with minimal friction: *“It shouldn’t take a hundred extra clicks, and I shouldn’t have to search for it — ideally it pops up, or I have to actively dismiss it. Sometimes you have to give people a little nudge to get them to read things“* (P7). Integration into existing clinical systems was considered essential; passive availability was broadly rejected.

### Phase 2: Prototype development

Qualitative findings were translated into a structured set of design requirements, encompassing user needs, design consequences, and resulting features, that guided iterative co-creation of the Clinician Model Card with physicians, designers, and data scientists. The full traceability chain, from interview code to user need, design requirement, prototype feature, and evaluation survey item, is documented in Supplementary Data 1. This chain reflects a systematic effort to ensure that each design decision had an explicit qualitative origin and a corresponding quantitative test. The prototype was built around a concrete clinical use case — a recurrent neural network–based CDSS that predicts sepsis probability over the next four hours using electronic health record data from the emergency department — to ensure evaluation in a realistic clinical context. However, not all expressed clinician preferences could be fully realized in this prototype iteration. Instance-level explainability, the most frequently mentioned unmet information need, requires integration at the CDSS level and cannot be delivered through documentation alone. This boundary condition is reflected in the design principles proposed.

The most fundamental departure from the Model Facts Label was the conversion from a PDF to an interactive digital interface^20^, designed for integration into a CDSS or hospital information system. This shift enabled structural and presentational changes that an unlayered format cannot support. Section ordering was revised to reflect the relevance hierarchy identified in Phase 1: *Uses & Directions* was elevated and reformatted as a scannable bullet list with an accompanying workflow flowchart; *Warnings* was repositioned higher with added iconography; and *Validation & Performance* was moved toward the end, consistent with its lower perceived clinical relevance. The *Mechanism* section was renamed *Data & Mechanism* and restructured into a tabular format. A persistent sidebar navigation panel and a condensed summary quick-start guide were added to support rapid orientation under time pressure.

To address the comprehensibility challenges identified in Phase 1, particularly around statistical terminology, interactive tooltips were embedded throughout the *Validation & Performance* section, providing plain-language explanations on demand without cluttering the primary interface, following Shneiderman’s principle of overview first, details on demand^22^. The tooltips translated conditional probabilities into natural frequencies and absolute terms, for example, explaining a PPV of 14% as “out of 100 patients flagged as positive, approximately 14 will actually have the condition”, to support the interpretation of probabilistic model outputs and reduce the cognitive demands of interpreting raw metric values^23^. The overall design adheres to WCAG 2.1 Level AA accessibility guidelines^24^.

The prototype was named the Clinician Model Card and is shown in Fig. 1; the full wireframe is available at: https://www.figma.com/proto/SHeuT3okLaxPMxs6bG7Jxj/clinician_model_card?node-id=2-238&t=HEhfgvHyiA80xad8-0&scaling=scale-down&content-scaling=fixed&page-id=0%3A1&starting-point-node-id=2%3A238. A video demonstration is provided in Supplementary Video 1.

**Fig. 1:**
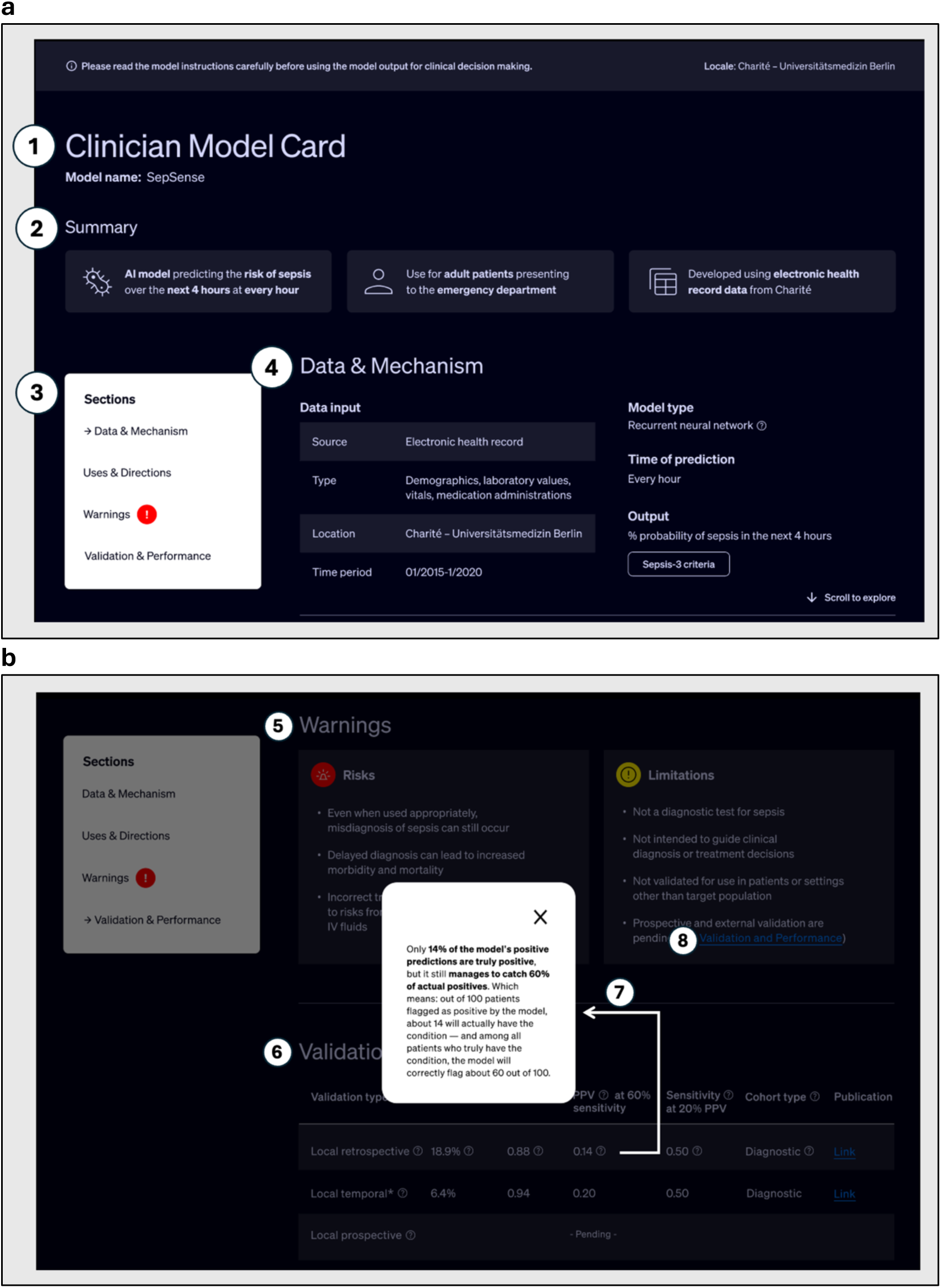
Clinician Model Card prototype interface. “SepSense” shown as exemplary use case. **a** Landing page. **b** *Warnings* and *Validation & Performance* sections with open interpretation tooltip. Annotated elements: (1) title and model name; (2) *Summary*: intended use, target population, data source; (3) persistent sidebar with click-to-jump and scroll-based navigation; (4) *Data & Mechanism*: tabular model inputs, type, and output; (5) *Warnings*: color-coded risks and limitations; (6) *Validation & Performance*: cohort-level metrics with embedded interpretation tooltips; (7) open tooltip with frequency-format PPV interpretation (background dimmed); (8) cross-link to related section. PPV, positive predictive value.

### Phase 3: Quantitative survey

#### Participants

Of 710 clinicians who opened the survey link, 254 initiated the survey. Two analytical samples are used throughout: 129 valid cases, defined as completing all demographic items including the AI literacy scale and responding to at least one Clinician Model Card item, and 109 complete cases who answered every non-free-text item. Unless otherwise noted, results are reported for the valid-case sample (n = 129); items with differential dropout are reported with their actual denominators. Median response time was 10.1 minutes (IQR 7.8–12.2). Mean AI literacy (SNAIL-TU; a 14-item validated scale of AI technical understanding^25,26^, 1–5) was 2.44 (SD = 1.07), and mean technical affinity (ATI-S; a 4-item validated scale^27^, 1–6) was 4.23 (SD = 1.10), indicating relatively low AI literacy and moderate technical affinity with substantial inter-individual variability. Participant characteristics are summarized in Table 1. The survey data, including open-text responses from 14 participants, are provided in Supplementary Data 2.

**Table 1:** Demographics of the 129 included survey respondents.

| Characteristics of Participants | n (%) |
| --- | --- |
| Valid case § | 129 (100) |
| Complete case * | 109 (84.5) |
| Finished survey + | 108 (83.7) |
| Response time, median (IQR), min | 10.1 (7.8–12.2) |
| Gender # |  |
| Female | 61 (47.3) |
| Male | 68 (52.7) |
| Age, median (IQR), years | 37 (30–47) |
| Professional experience, median (IQR), years | 8 (3–17) |
| Specialty |  |
| Internal medicine | 62 (48.1) |
| Anesthesiology | 30 (23.3) |
| Other \$ | 37 (28.7) |
| Experience with AI-based CDSS |  |
| Yes | 30 (23.3) |
| No | 99 (76.7) |
| Setting |  |
| Academic medical center | 105 (81.4) |
| Non-academic medical center | 16 (12.4) |
| Other | 8 (6.2) |
| SNAIL – Technical understanding, mean (SD) | 2.44 (1.07) |
| ATI-S, mean (SD, α) | 4.23 (1.1, 0.82) |

§ All demographic questions answered, incl. SNAIL, and at least one question on the Clinician Model Card answered. \* Every question answered excluding free-text answers. + Reached last page. # Options included female, male, and non-binary. \$ Includes specialties with fewer than 8 respondents or respondents who selected 'Other'. CDSS, clinical decision support system; IQR, Interquartile range; SD, standard deviation; SNAIL, Scale for the Assessment of Non-Experts' AI Literacy; ATI-S, Affinity for Technology Interaction Short Scale.

#### Objective comprehensibility

Four single-best-answer items assessed retention of information from the Clinician Model Card after a familiarization period in which participants were instructed to read all content carefully, scroll through the card, and interact with the available explanatory aids, particularly in the *Validation & Performance* section; the card was not accessible during this section. The mean score (Qn-C0.1–Qn-C0.4) was 2.26 of 4 possible points (56.4%). Item-level performance revealed a pronounced split: the item assessing intended clinical use, a non-statistical question, was answered correctly by 87.6% (Qn-C0.3). By contrast, items requiring interpretation of statistical metrics were answered correctly by fewer than half of respondents: AUROC interpretation 42.6% (Qn-C0.1), stage of validation 41.9% (Qn-C0.2), and PPV interpretation 53.5% (Qn-C0.4). This pattern of near-ceiling performance on clinical content and near-chance performance on statistical content despite the availability of explanatory tooltips during the familiarization phase is further examined in the subgroup analysis reported below.

#### Subjective comprehensibility

Perceived comprehensibility was high for most sections: *Summary* (74%, 92/124), *Warnings* (74%, 90/122), *Uses & Directions* (65%, 80/124), and *Data & Mechanism* (60%, 73/121) were each considered well understood by a clear majority (Fig. 2a). *Validation & Performance* was the sole exception: only 32% (39/123) agreed they understood it well, while 46% (56/123) actively disagreed — the only section where disagreement exceeded agreement, and a pattern that persisted despite the addition of interactive tooltips and plain-language explanations.

**Fig. 2:**
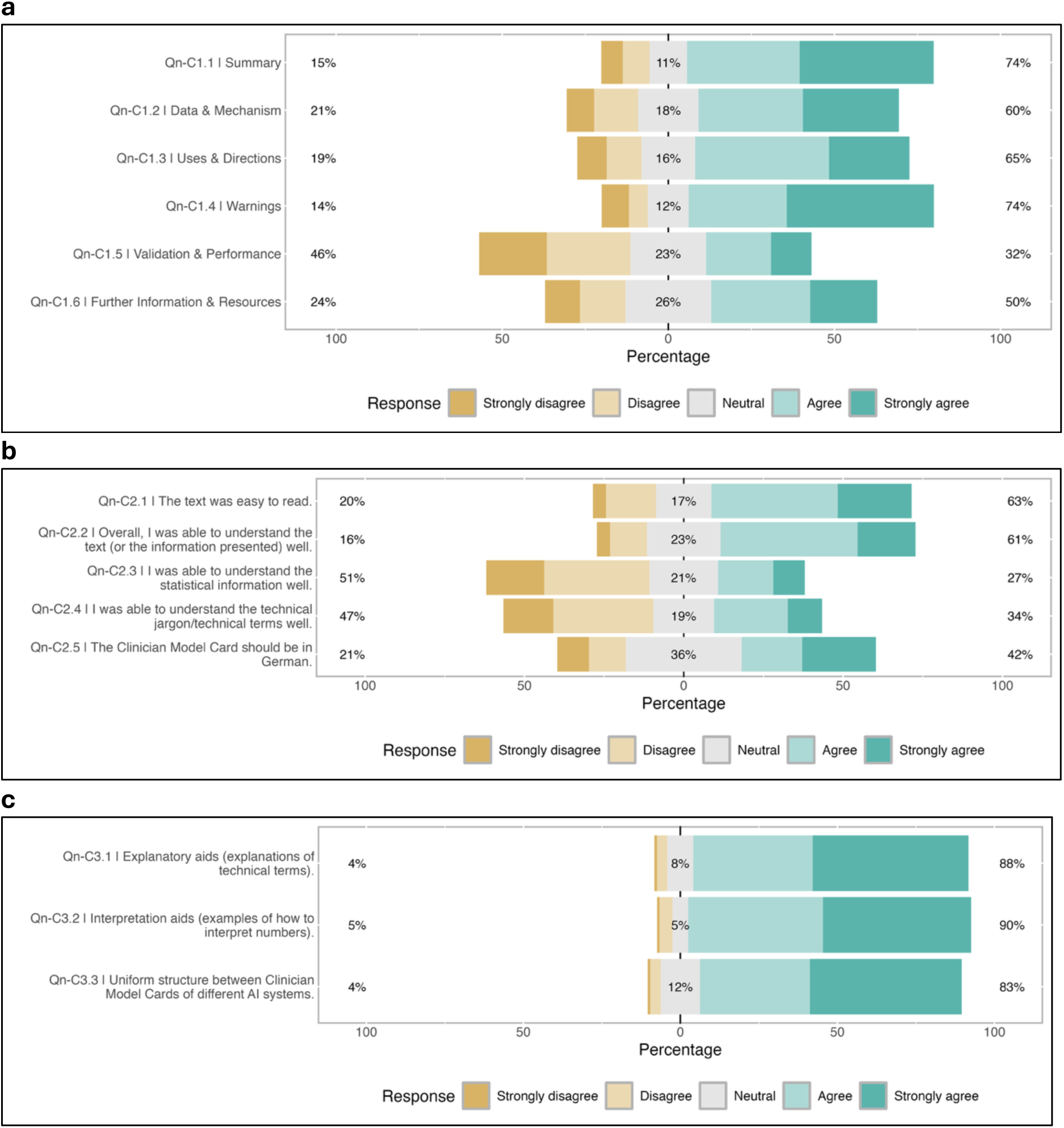
Perceived comprehensibility of the Clinician Model Card. **a** Comprehensibility ratings for individual sections of the Clinician Model Card. **b** Agreement with statements assessing overall comprehensibility of the Clinician Model Card. **c** Factors perceived to improve comprehensibility of the Clinician Model Card.

Overall, 63% found the card easy to read (76/121) and 61% felt they understood the information well (74/121); however, comprehensibility of statistical content specifically was limited, with only 27% agreeing they understood the statistical information (33/121) and 34% the technical terminology (41/121) (Fig. 2b). The interpretive enhancements were nonetheless broadly valued: 88% found explanatory aids helpful (106/121) and 90% valued interpretation aids (109/121), indicating that clinicians recognized the aids as useful even when comprehension remained incomplete (Fig. 2c).

#### Relevance

Two-thirds of participants considered the Clinician Model Card relevant to clinical decision-making (66%, 75/113; Qn-R1.2). Views on completeness were more divided: 39% felt the content was sufficient while 46% were neutral (Qn-R1.1); 44% were satisfied with the level of detail while 34% would have preferred more (50/113 and 38/112 respectively; Qn-R1.3, Qn-R1.4) (Fig. 3a). The most frequently requested additional information was subgroup-specific performance — how reliably the system performs across different patient populations — endorsed by 83% (95/115; Qn-R2.3), followed by model explainability (64%, 74/115; Qn-R2.4), evidence and validation (57%, 66/115; Qn-R2.2), and training data characteristics (56%, 64/115; Qn-R2.1) (Fig. 3b).

**Fig. 3:**
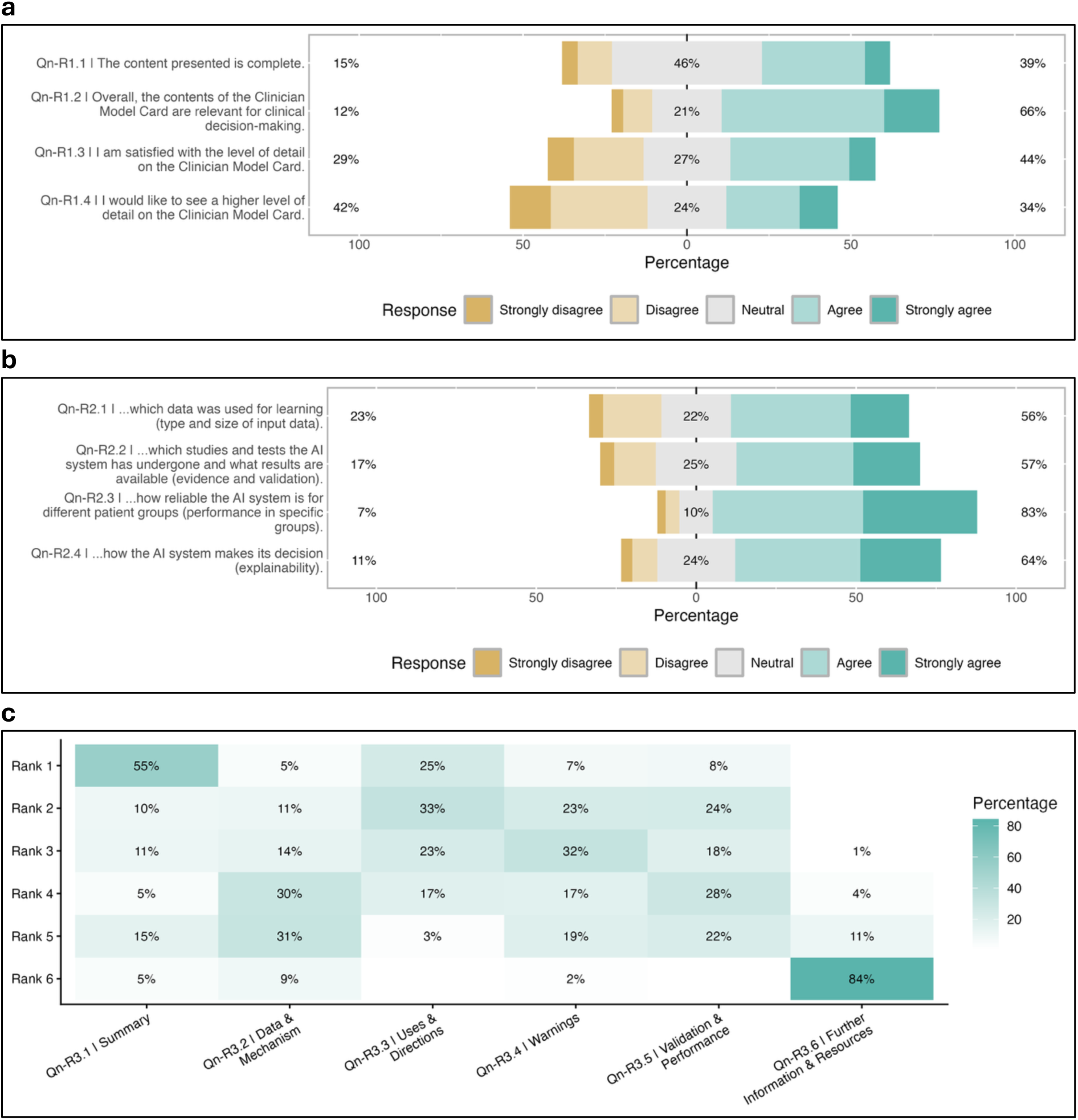
Relevance of the Clinician Model Card for clinical decision-making. **a** Physician ratings of the overall relevance, usefulness, level of detail, and completeness of the Clinician Model Card. **b** Types of additional information physicians considered useful to include in the Clinician Model Card (question prompt: ‘I would like more information within the Clinician Model Card about…’). **c** Heatmap of physician rankings of the six Clinician Model Card sections by perceived clinical importance, from most to least important.

Section rankings closely replicated the card sorting results from Phase 1 interviews, providing cross-phase validation of the relevance hierarchy. *Summary* and *Uses & Directions* were ranked first and second (median ranks 1 and 2; Qn-R3.1, Qn-R3.3); *Warnings* ranked third (median rank 3; Qn-R3.4), higher than its position in the original Model Facts Label; *Validation & Performance* ranked fourth (median rank 3.5; Qn-R3.5); and *Information & Resources* was ranked last most often (median rank 6; Qn-R3.6). A heatmap of rankings is shown in Fig. 3c.

#### Accessibility

Participants strongly preferred active, workflow-integrated access over passive availability. The vast majority agreed the card should be accessible during AI system use, that the CDSS should proactively prompt its use, and that contextual alerts should direct clinicians to the card when there is risk of inappropriate use — for example, when a model validated in adults is applied to a pediatric patient (84%, 84%, and 85% respectively; 91/108, 91/108, 93/108; Qn-A1.2, Qn-A1.3, Qn-A1.4). Three-quarters agreed the card should be provided before first use (76%, 82/108; Qn-A1.1) (Fig. 4a). For delivery channel, direct integration within the CDSS or hospital information system was the clear preference (93%, 99/107; Qn-A2.1), with the intranet or clinical portal as a secondary option (77%, 82/107; Qn-A2.3); email and print formats were rejected by majorities of 55% and 71% respectively (Qn-A2.2, Qn-A2.4).

**Fig. 4:**
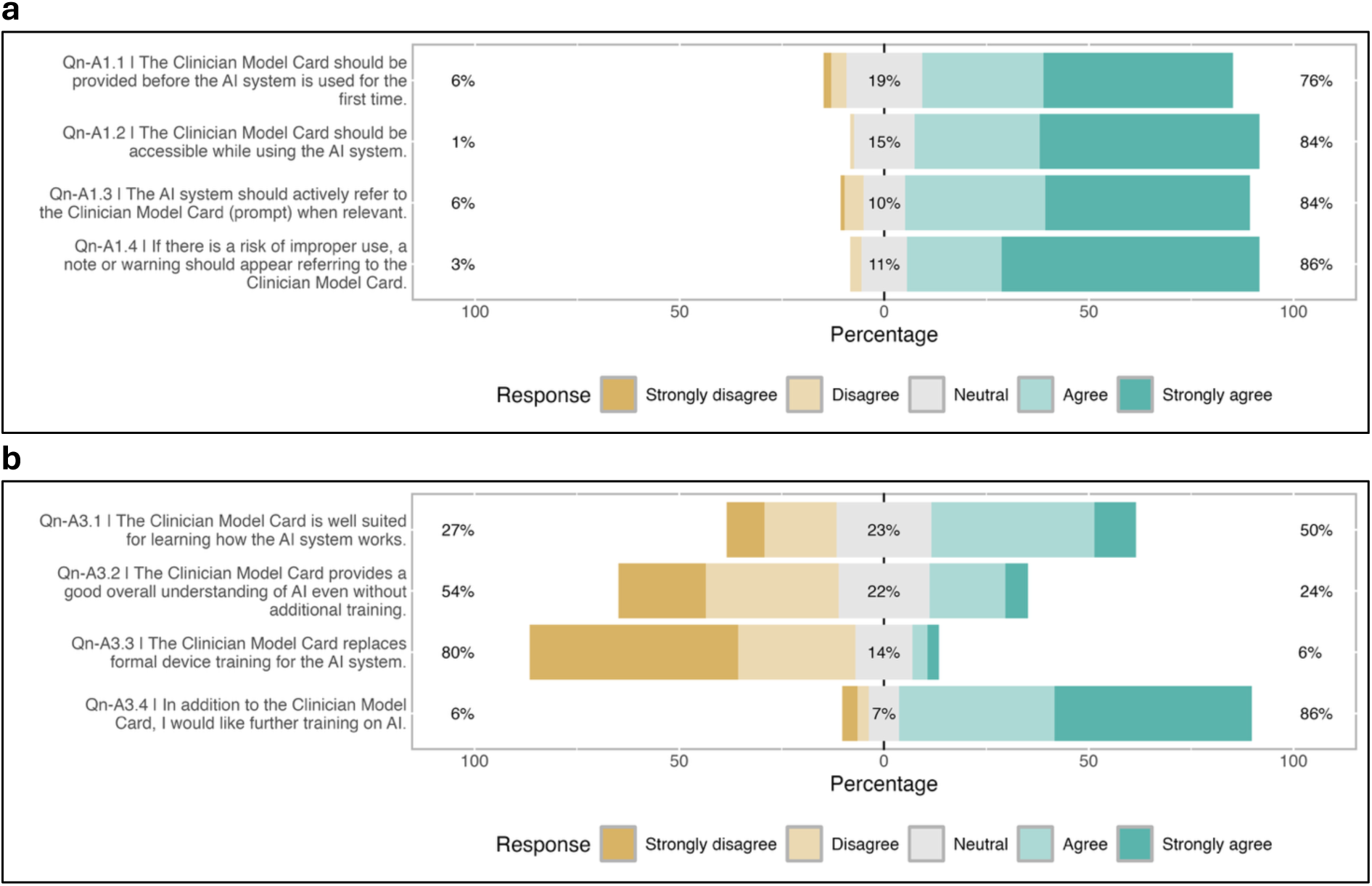
Accessibility of the Clinician Model Card. **a** Physician preferences for when and how the Clinician Model Card should be accessible during clinical decision-making. **b** Perceived role of the Clinician Model Card in supporting AI literacy and training, including the need for additional training beyond the card.

Participants drew a clear boundary between documentation tool and AI training: 86% indicated a desire for additional AI training beyond the card (93/108; Qn-A3.4), 80% agreed it should not replace device-specific onboarding (86/108; Qn-A3.3), and only half felt the card adequately supported understanding of how the AI system works (50%, 54/108; Qn-A3.1) (Fig. 4b).

### Exploratory subgroup analysis

Spearman rank correlations between SNAIL-TU and ATI-S scores and comprehensibility outcomes were conducted to examine whether AI literacy and technical affinity is associated with comprehension of the Clinician Model Card. AI literacy showed a large positive correlation with overall objective comprehension (ρ = 0.59, p < 0.001) and all individual items, with associations strongest for the most technical items — AUROC interpretation (ρ = 0.52) and stage of validation (ρ = 0.39) — and weakest for the clinically framed item (ρ = 0.19; all p < 0.05). This gradient directly mirrors the item-level performance pattern reported above. Subjective comprehensibility of statistical content showed a similar pattern (ρ = 0.49 for both Qn-C2.3 and Qn-C2.4, p < 0.001). Technical affinity showed weaker but consistent associations across the same outcomes (ρ = 0.18–0.44).

For relevance, higher AI literacy and technical affinity were each associated with greater perceived relevance of the card for clinical decision-making (ρ = 0.31, p < 0.001 and ρ = 0.26, p = 0.008 respectively; Qn-R1.2), suggesting that technical understanding of AI increases rather than reduces the perceived value of structured documentation. Desire for more detail showed no significant association with either predictor (SNAIL-TU: ρ = 0.18, p = 0.054; ATI-S: ρ = 0.14, p = 0.15; Qn-R1.4), indicating that demand for additional content was broadly distributed across literacy levels.

## Discussion

The Clinician Model Card demonstrates both the potential and the limits of documentation-led approaches to clinical AI transparency. Grounded in empirically derived clinician needs and evaluated in a national sample of 129 physicians, the tool was well-received in most respects: the majority of participants found it readable and clinically relevant, strongly preferred active workflow integration over passive availability, and replicated the relevance hierarchy identified in Phase 1 — with *Summary* and *Uses & Directions* ranked highest and *Validation & Performance* lowest in both the interview card sort and the survey. This cross-phase convergence between a small, single-institution, qualitative sample and a national, multi-specialty survey suggests that the priorities we identified are not idiosyncratic, but rather reflect a broader, consistent pattern of clinician information needs, at least regarding relevance and accessibility. However, comprehensibility, particularly with regard to statistical content, proved to be more resistant to design intervention alone.

Yet the central finding of the study is a tension that documentation design alone has not resolved: despite targeted interventions (interactive tooltips, plain-language explanations, and deliberate repositioning of statistical content) comprehensibility of the *Validation & Performance* section remained poor. Only 32% of respondents agreed they understood it well, while 46% actively disagreed, and fewer than half correctly interpreted AUROC or PPV in the objective comprehension items. This pattern is consistent with prior investigations into clinicians’ understanding of AI performance metrics^19^ and is reinforced by the subgroup analysis: AI literacy (SNAIL-TU) was the strongest predictor of objective comprehension (ρ = 0.59), with the steepest gradient precisely on the most technical items, suggesting the barrier is not one of information access alone but of the prior knowledge required to engage with it^3,18^.

This finding is deepened by the relevance data: participants ranked *Validation & Performance* last among all sections — the section most critical for appropriate use — reflecting a well-documented tendency for unfamiliar concepts to be underweighted in clinical judgment^28,29^. Positioning *Validation & Performance* toward the end of the card may reflect clinician relevance rankings, but this ordering creates a dead angle precisely where vigilance is most needed. The justification is pragmatic rather than principled: a card that loses the reader before reaching statistical content fails entirely. This trade-off should be made explicit, and contextual triggers surfacing performance information at decision-critical moments are essential to compensate.

That this gap persists even when explanatory aids are available was anticipated by at least some participants. As one survey participant noted: *“Statistics is complex but important. Someone who didn’t develop this themselves won’t understand everything immediately, even with the pop-ups. Links to external explanations would be helpful, especially since these would apply to all models“* (CASE49). The implication is not to remove or further deprioritize this content, but to recognize that documentation design is necessary and not sufficient: meaningful AI transparency requires parallel investment in statistical and AI literacy, integrated into medical curricula at undergraduate and postgraduate levels^30–32^ and in ongoing training for practicing clinicians.

The strong and consistent preference for proactive, workflow-integrated access — 93% preferred CDSS or hospital information system delivery, 84% wanted the CDSS to actively prompt use, and 85% supported contextual alerts for potential misuse — underscores that accessibility is not a question of format alone. Two distinct contexts of use shaped participants’ access preferences: an onboarding phase requiring comprehensive engagement, and on-demand reference during routine care, triggered by unexpected or contradictory results and requiring rapid, targeted access. A layered interface^17^ — concise by default, with expandable detail on demand^22^ — is designed to serve both contexts within a single artifact. The interactive architecture of the Clinician Model Card builds on prior work by Crisan et al.^20^, who demonstrated the feasibility and value of human-centered, interactive model documentation — an approach our findings now evaluate empirically in a clinical context. A Clinician Model Card that must be sought out will not be consulted, especially not under time pressure. The tool should be treated as a functional component of the CDSS: surfaced contextually, triggered proactively, and embedded where clinical decisions are made, rather than as a document that happens to be available nearby.

A related but distinct boundary condition emerged with equal clarity: 86% of respondents desired additional AI training beyond the card, and only half felt it adequately supported understanding of how the AI system works. The Clinician Model Card is designed to support informed use at the point of care: to help clinicians apply an AI system appropriately, recognize its limitations, and avoid misuse. It is not a training resource, and conflating these roles risks both over-reliance on documentation as an educational substitute and under-investment in the training infrastructure that is genuinely needed.

Sixty-four percent of respondents wanted to understand how the AI system arrives at its predictions — not aggregate validation statistics, but the rationale behind *this* prediction for *this* patient. This clinically intuitive desire for instance-level explainability cannot be addressed through model card documentation, regardless of how well designed. Explanations of individual predictions require integration of explainable AI methods, such as attention maps, SHAP values, or natural language explanations, directly within the CDSS interface, and these come with their own interpretive challenges and implementation requirements that warrant dedicated investigation^33,34^. Documentation and explainability are complementary but structurally distinct interventions.

The highest-ranked unmet information need was subgroup-specific performance (83%, 95/115): how reliably the system performs across different patient populations. This is clinically intuitive and practically important (e.g., a model validated primarily in one demographic may perform poorly in another) yet subgroup performance remains among the most inconsistently reported aspects of AI model documentation^35^. Disaggregated performance reporting should be treated as a minimum standard for clinical AI, surfaced prominently within the model card rather than buried in technical appendices. Our findings add to a growing body of evidence that the field has not yet established adequate norms for equity-relevant documentation^35,36^, and that clinicians themselves recognize and demand this information when given the opportunity to express it.

The EU AI Act and MDR represent meaningful steps toward accountable clinical AI, mandating that high-risk systems be accompanied by clear, accessible documentation for end users^8,37^. Yet our findings illustrate that compliance with transparency requirements does not guarantee utility in practice. The Model Facts Label was already a thoughtful clinician-oriented framework, yet Phase 1 interviews revealed consistent shortcomings in relevance, comprehensibility, and accessibility. The Clinician Model Card addresses many of these, but the persistent comprehensibility gap demonstrates that well-intentioned documentation can still fail its intended audience. The design principles reported here offer a concrete operationalization of what the EU AI Act requires but does not specify, and they do so on an empirical rather than a normative basis. Actionable transparency for clinicians requires iterative co-design and empirical evaluation, not documentation produced for regulatory purposes and handed to users untested.

The principles that emerge from this study are not specific to the Clinician Model Card or to sepsis prediction. They reflect structural features of how clinicians engage with AI documentation: how they weigh relevance, where they encounter comprehension barriers, and under what conditions they will or will not consult a transparency tool. Features that are likely to generalize across CDSS contexts. The findings of this study converge on a set of empirically derived principles for clinician-centered AI model documentation (Fig. 5), organized around relevance, comprehensibility, and accessibility, with a fourth category capturing what documentation alone cannot achieve. A complete mapping of qualitative codes and quantitative survey items to each principle is provided in Supplementary Data 1.

**Fig. 5.**
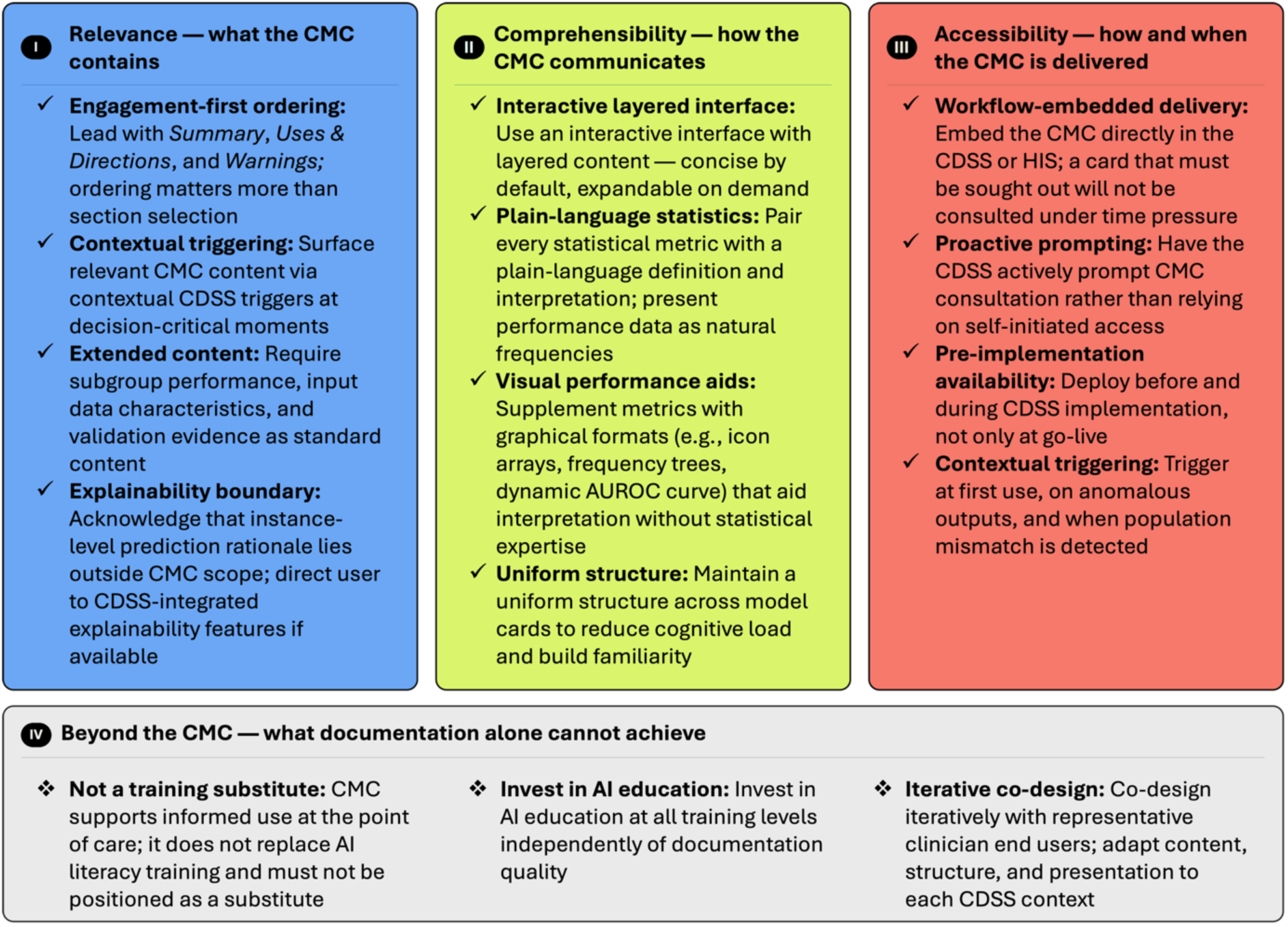
Empirically derived design principles for clinician-centered AI model documentation, organized by Relevance, Comprehensibility, and Accessibility. AUROC, area under the receiver operating characteristic curve; CDSS, clinical decision support system; CMC, Clinician Model Card; HIS, hospital information system.

This study has several limitations. Survey recruitment relied on convenience and snowball sampling, introducing potential selection bias toward clinicians with greater interest in digital health; the predominance of academic medical center participants (81%) may mean AI literacy and technological affinity were above average for the broader physician population. The prototype was evaluated in isolation rather than within a functioning CDSS, so findings reflect perceptions of a standalone artifact rather than behavior in real clinical use. Some item arrays showed limited response variance, raising the possibility of acquiescence bias in a self-constructed instrument. As an exploratory study without confirmatory hypotheses, no power calculation was performed. Finally, SepSense is a high-acuity, time-pressured use case; design requirements may differ for lower-stakes or less time-sensitive CDSS applications, and generalizability across clinical contexts should be tested empirically.

This was a first-generation prototype, and the findings point clearly toward what the next generation should address. More intuitive visualization of model performance, particularly for subgroup-specific metrics and for statistical content that currently eludes most clinicians, remains the most pressing design challenge. Richer reporting of training data characteristics and validation evidence was also frequently requested and warrants inclusion in future iterations. Most importantly, the central question this study could not answer is whether access to a Clinician Model Card actually improves clinical decision-making. Prospective evaluation of the tool embedded within a functioning AI-CDSS, assessing whether structured model documentation influences how clinicians act on AI outputs, and ultimately whether it contributes to safer care — is the necessary next step.

The Clinician Model Card demonstrates that clinician-centered, interactive AI documentation is feasible, valued by clinicians, and capable of improving upon its predecessors in relevance, structure, and accessibility. The persistent comprehensibility gap around statistical performance metrics, however, confirms that no documentation tool can substitute for the educational and implementation infrastructure that responsible clinical AI deployment requires. Effective transparency is not a document — it is a system property, emerging from the combination of thoughtful design, workflow integration, and investment in clinician AI literacy.

## Methods

### Study design

We used a sequential exploratory mixed-methods design following the digital tool development variant^21^, comprising three phases: (1) qualitative exploration via semi-structured physician interviews, (2) prototype development informed by qualitative findings, and (3) quantitative evaluation of the prototype via a physician survey.

### Ethical approval and consent

Ethics approval was obtained from the institutional review board of Charité – Universitätsmedizin Berlin (EA4/140/24). All participants provided informed consent prior to participation.

### Phase 1: Qualitative interviews

The goal of Phase 1 was to elicit clinician information needs, comprehension barriers, and accessibility requirements with respect to AI model documentation, and to translate these into design requirements for the documentation tool prototype.

#### Participants and recruitment

Twelve physicians from internal medicine and anesthesiology at Charité – Universitätsmedizin Berlin were recruited between December 2024 and January 2025 using purposive sampling to ensure variation in clinical specialty, care setting, seniority, and gender. Sampling was informed by the study’s CDSS use case (AI-based sepsis prediction in the emergency department) and targeted physicians familiar with this clinical scenario. Recruitment continued until data saturation was reached, with an a priori target of 10–15 interviews.

#### Interview design and procedure

The semi-structured interview guide was adapted from Crisan et al.^20^ and extended to integrate the transparency dimensions specified in Article 13 of the EU AI Act — relevance, comprehensibility, and accessibility of instructions for high-risk AI systems. A digital Miro board (Miro DACH GmbH, Berlin, Germany) was used to present the Model Facts Label^15^ and to facilitate a card sorting task, in which participants ranked its eight predefined sections by perceived clinical relevance. The Miro board also included visual and interactive design inspirations drawn from Crisan et al.^20^, such as the model performance panel and the ‘Robustness Gym Report’, to elicit preferences regarding visualization and interactivity. A structured discussion of each section’s relevance, comprehensibility, and accessibility followed, including a mapping exercise in which participants assigned EU AI Act transparency requirements to card sections. At the conclusion of each interview, participants were asked whether any information they considered important was missing. Participants were invited to think aloud throughout. Prior to data collection, the interview guide was refined through pre-tests with four physicians. Interviews were conducted by one researcher (LAMS) in German, either face-to-face or via Microsoft Teams, audio-recorded, and transcribed verbatim using Whisper (OpenAI, San Francisco, CA, USA) with manual correction. The interview structure is visualized in Fig. 6; the full interview guide and Miro board are provided in Supplementary Methods 1 and 2.

**Fig. 6:**
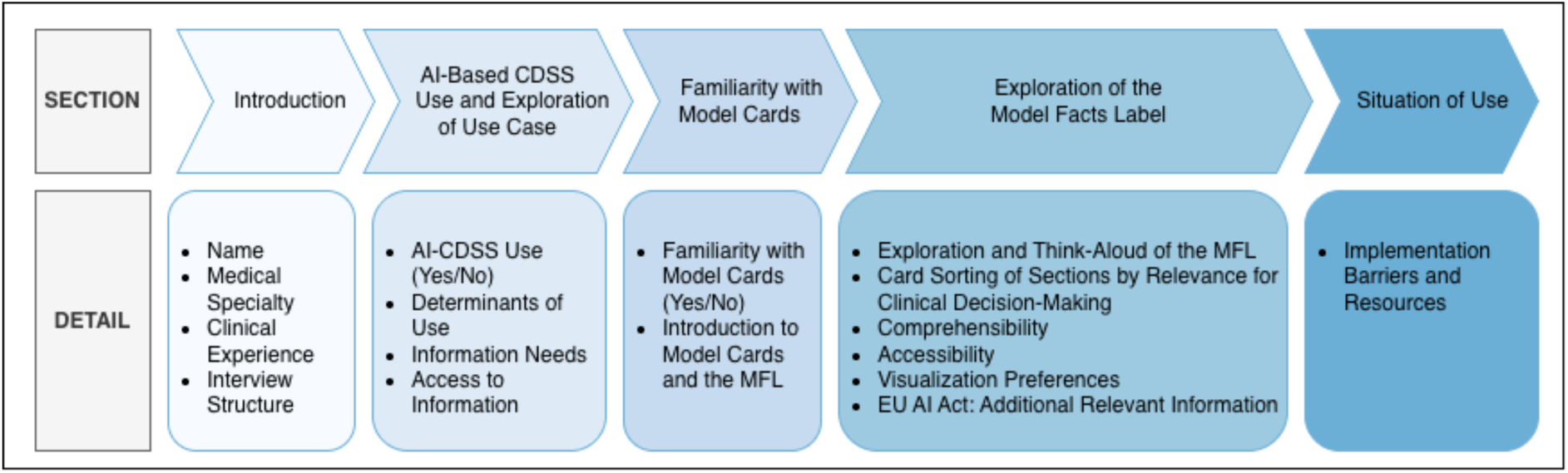
Interview structure and sequence. The interview comprised five sections: introduction and participant characteristics, AI-based CDSS use and exploration of the clinical use case, familiarity with Model Cards, exploration of the Model Facts Label, and situation of use. The Model Facts Label exploration included a think-aloud protocol, card sorting of sections by perceived clinical relevance, and structured discussion of comprehensibility, accessibility, visualization preferences, and EU AI Act-relevant information. The final section addressed implementation barriers and enablers. CDSS, clinical decision support system; EU AI Act, European Union Artificial Intelligence Act; MFL, Model Facts Label.

#### Analysis

Transcripts were analyzed using qualitative content analysis^38^ in MaxQDA (VERBI GmbH, Berlin, Germany), following a deductive-inductive approach^39^. Two researchers independently coded all transcripts; codes were then discussed and harmonized across the research team to produce a final coding framework with clearly defined categories, which was applied consistently across the dataset. Intercoder agreement is reported in Supplementary Methods 3. Card sorting data were analyzed by assigning each section a score of 1–8 per participant based on placement order; scores were summed across participants and inverse-weighted to yield a final section ranking. Data saturation was assessed through ongoing team reflection and confirmed when no new categories emerged across successive interviews.

### Phase 2: Prototype development

The goal of Phase 2 was to translate qualitative findings into a functional, clinician-centered prototype for clinical AI documentation through a structured rapid prototyping process comprising four stages: analysis, definition, design, and test and optimization. Qualitative findings were synthesized using a structured insight-to-decision mapping approach, translating codes into user needs and design consequences (Supplementary Data 1). This synthesis informed a visual co-creation process involving physicians, designers, and data scientists. In this process, design decisions were developed collaboratively and made directly visible, rather than solely being derived abstractly from interview summaries. Then, low-fidelity wireframes were iteratively developed and revised through physician feedback focused on navigation structure, language clarity, and layout, then progressively refined into a high-fidelity interactive prototype built in Figma (Figma, Inc., San Francisco, CA, USA). The visual design underwent multiple iterations informed by usability testing with physicians, with findings from each round incorporated before retesting. The factual content of the Model Facts Label was preserved and adapted to the local implementation context throughout. The resulting tool was named the Clinician Model Card.

### Phase 3: Quantitative evaluation

The goal of Phase 3 was to evaluate the Clinician Model Card prototype in a national sample of physicians, assessing its comprehensibility, clinical relevance, and accessibility as primary outcomes.

#### Participants and recruitment

Physicians were recruited across medical centers in Germany between October 2025 and January 2026 via personal outreach, morning round presentations, institutional email lists, flyers, medical association networks, and peer distribution. Inclusion required completion of all demographic items including the SNAIL-TU scale, and a response to at least one Clinician Model Card evaluation item. A complete case sample (defined as answering every non-free-text survey item) was used for analyses requiring full item coverage. As this was an exploratory study, no formal power calculation was performed; a minimum of 100 analyzable responses was set as a pragmatic threshold.

#### Survey instrument

The online survey was administered in German via SoSci Survey (SoSci Survey GmbH, Munich, Germany) and structured into six sections: introduction and study information, demographics, AI literacy, Clinician Model Card evaluation (including guided prototype exploration and items assessing comprehension, relevance, and accessibility), technical affinity, and open feedback. Participants were presented with a standardized clinical scenario set in a hospital emergency department, in which a fictive AI-based CDSS (“SepSense”) predicted the probability of sepsis over the next four hours. The Figma prototype was embedded directly into the survey via HTML; progression to the evaluation section was blocked for 45 seconds to ensure a minimum familiarization period. Survey items were derived directly from the design requirements established in Phase 2, with items targeting a specific feature or qualitative finding (for item-level traceability from qualitative findings to quantitative evaluation refer to Supplementary Data 1). Agreement with survey statements was measured on five-point Likert scales; perceived relevance was assessed via drag-and-drop ranking from most to least important. AI literacy was assessed using the technical understanding subdomain of the SNAIL scale^25,26^ and technical affinity using the ATI-S^27^, both in validated German versions. Cognitive interviews were conducted prior to data collection to assess survey comprehensibility. The survey structure is shown in Fig. 7. An English version of the survey is available at https://survey.charite.de/CMC_ENG/ (password: CMC).

**Fig. 7:**
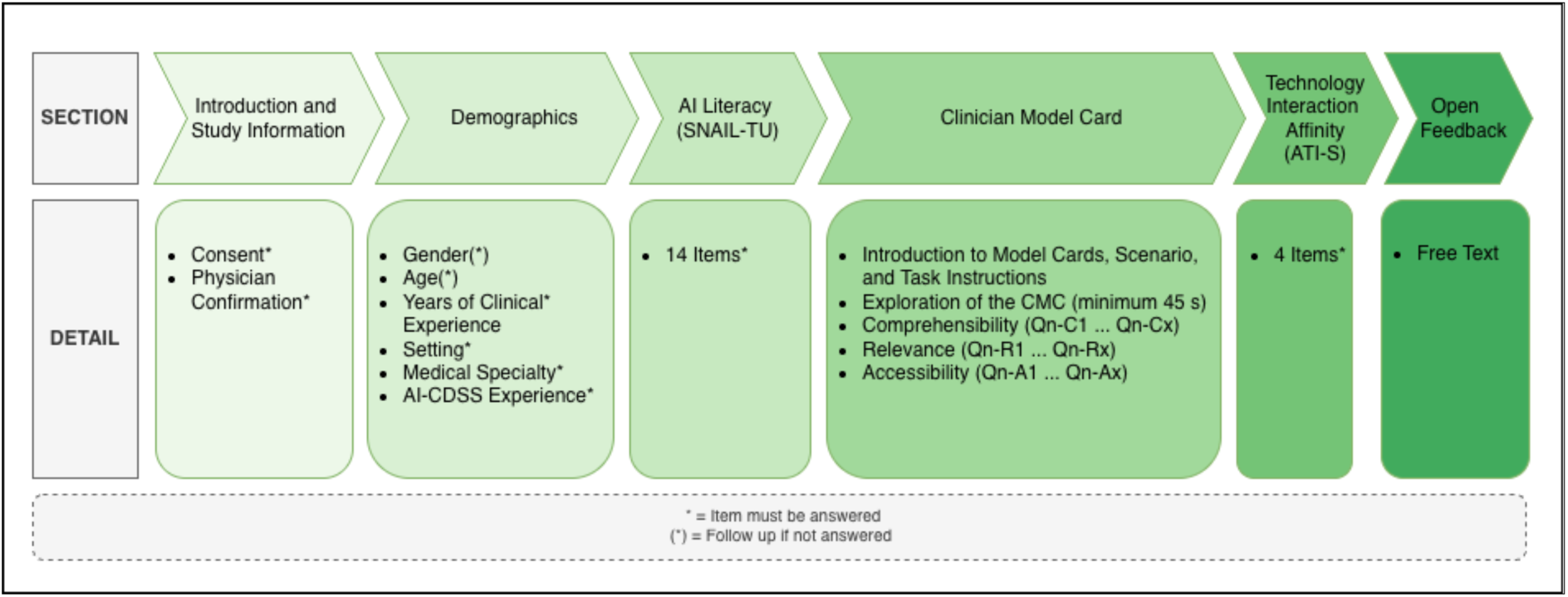
Survey structure and sequence. The survey comprised six sections: introduction and study information, demographics, AI literacy, Clinician Model Card evaluation, technical affinity, and open feedback. The Clinician Model Card evaluation section included a standardized clinical scenario, a minimum 45-second prototype exploration period, and items assessing comprehensibility (Qn-C), relevance (Qn-R), and accessibility (Qn-A). Asterisks indicate mandatory items; items marked (*) were followed by a mandatory response if left blank. ATI-S, Affinity for Technology Interaction Short Scale; CDSS, clinical decision support system; CMC, Clinician Model Card; SNAIL-TU, Scale for the Assessment of Non-Experts’ AI Literacy, Technical Understanding subdomain.

#### Statistical analysis

Descriptive statistics are reported for all participant characteristics and survey response metrics. Individual Likert items are summarized as proportion agreement (combining “agree” and “strongly agree”) and proportion disagreement (combining “disagree” and “strongly disagree”), reported as n/N (%); full descriptive statistics including bootstrapped 95% confidence intervals (10,000 iterations) are provided in Supplementary Data 4. Comprehension items are reported as proportion correct per item and mean total score. Section rankings are summarized as median rank. AI literacy (SNAIL-TU) and technical affinity (ATI-S) are reported as mean and standard deviation; Cronbach’s alpha was computed for the ATI-S. Likert responses are visualized as diverging stacked bar charts and rankings as a heatmap. Spearman rank correlations were used to examine associations between AI literacy, technical affinity, and comprehensibility and relevance outcomes. All analyses were conducted in R version 4.4.1.

## Acknowledgments

We thank the participants who shared their perspectives during our interviews and surveys. This work builds upon the Model Facts Label developed by Sendak et al. and was further inspired by the research of Crisan et al. We also appreciate the constructive partnership with denkwerk GmbH in developing the Clinician Model Card. Lastly, we thank Dr. Matthias Laupichler for his valuable advice regarding the use of SNAIL.

## Data availability

Supplementary materials (Supplementary Tables 1–3, Supplementary Methods 1–3, Supplementary Data 1–4, and Supplementary Video 1) are provided on OSF (https://osf.io/7w8hr). Interview transcripts cannot be shared publicly as participants did not consent to data release beyond the research team, in accordance with the ethics approval granted by the Charité – Universitätsmedizin Berlin institutional review board (EA4/140/24).

## Contributions

LAMS: Conceptualization; Methodology; Investigation; Data Curation; Formal Analysis; Visualization; Writing – Original Draft; Project Administration. ZK: Formal Analysis (qualitative coding; quantitative verification); Writing – Review & Editing. LM, CMB, FB: Supervision; Writing – Review & Editing. NF, EW, OF, JM, MM, AM, SG: Writing – Review & Editing.

